# Clinical features and outcomes of 2019 novel coronavirus–infected patients with cardiac injury

**DOI:** 10.1101/2020.03.11.20030957

**Authors:** Youbin Liu, Jinglong Li, Dehui Liu, Huafeng Song, Chunlin Chen, Mingfang Lv, Xing Pei, Zhihui Qin, Zhongwei Hu

**Author notes:** These authors are equal to the work (Youbin Liu, Jinglong Li, Dehui Liu, Huafeng Song, Chunlin Chen). Corresponding author: Youbin Liu; Department of Cardiology, Guangzhou Eighth People’s Hospital;postal address: NO 8 huaying road, Guangzhou city, Guangdong provice, PR China, Zhongwei Hu; Department of Internal medicine, Guangzhou Eighth People’s Hospital; postal address: NO 8 huaying road, Guangzhou city, Guangdong provice, PR China.

## Abstract

**Aims:** To explore the epidemiological and clinical features of 2019 novel coronavirus(2019-nCoV)-infected patients with cardiac injury.

**Methods and results:** Data were collected from patients’ medical records, and we defined cardiac injury according to cardiac biomarker troponin I level > 0.03 μg/L. Among the 291 patients, 15 (5.2%) showed evidence of cardiac injury. Of 15 hospitalized patients with cardiac injury, the median age was 65 years, and 11/15 (73.3%) were men. Underlying cardiovascular diseases in some patients were hypertension (n=7, 46.7%), coronary heart disease (n=3, 20%) and diabetes (n=3, 20%). The most common symptoms at illness onset in patients with cardiac injury were fever (n=11, 73.3%), cough (n=7, 46.7%), headache or fatigue (n=5, 33.3%) and dyspnea (n=4, 26.7%). These patients had higher systolic pressures, white blood cell count, neutrophil count, troponin I, brain natriuretic peptide, D-dimer and lower lymphocyte count, and platelet count, compared with patients without cardiac injury, respectively. Bilateral infiltrates on chest X-ray and elevated C-reactive protein occurred in all patients with cardiac injury. Compared with patients without cardiac injury, patients with cardiac injury were more likely to develop acute respiratory distress syndrome, and receive mechanical ventilation, continuous renal replacement therapy, extracorporeal membrane oxygenation and vasopressor therapy and be admitted to the intensive care unit.

**Conclusion:** Cardiac injury is a common condition among patients infected with 2019-nCoV. Compared with patients without cardiac injury, the clinical outcomes of patients with cardiac injury are relatively worse.

## Introduction

The 2019 novel coronavirus (2019-nCoV), a new fatal virus that emerged at the end of 2019, remains prevalent worldwide, especially in mainland China. As of March 11, 2020, at least 80969 cases have been diagnosed across mainland China, and 3162 people have died secondary to infection with this virus^1^. Because there is no specific drug therapy, the virus has caused public panic and great concerns globally. Although we know that infection with 2019-nCoV can lead to pneumonia, acute renal injury, acute respiratory distress syndrome (ARDS), and shock, we still know little about cardiac injury after infection with 2019-nCoV^2^.

Some viral infections are associated with cardiac injury^3^. Previous studies have indicated that cardiac damage by coronavirus is relatively mild, even with the most fatal coronaviruses, middle east respiratory syndrome and sudden acute respiratory syndrome^4, 5^. However, as a new coronavirus, we do not know if 2019-nCoV is harmful to the heart; the results from current studies are inconsistent. Chinese scientists have found no obvious damage to heart tissue caused by 2019-nCoV, according to autopsy results ^6^; however, other studies have reported that patients infected with 2019-nCoV often suffered cardiac injury ^7 8 9^. The relationship between cardiac injury and the risk of death among people infected with 2019-nCoV has not been clarified. Research is urgently needed to explore the clinical features and outcomes of 2019-nCoV-infected patients with cardiac injury.

Our study summarized the clinical characteristics of 2019-nCoV-infected patients with cardiac injury to provide insight into the prevention and treatment of heart disease, in these patients.

## Material and methods

We performed a retrospective study of the clinical characteristics of confirmed corona virus disease 2019(COVID-19)cases from 10 January 2020 to 24 February 2020. We identified 291 patients with confirmed 2019-nCoV infections according to laboratory testing after admission to hospital. The definitions of confirmed human infection with 2019-nCoV are based on the World Health Organization interim guidelines^10^. Only patients with a laboratory-confirmed infection were included in the present analysis. This study complies with the Declaration of Helsinki and was approved by the ethics commissions of the Guangzhou Eighth people’s hospital, with a waiver of informed consent.

The research team of the Department of Cardiology, Guangzhou Eighth People’s Hospital analysed patients’ medical records. Epidemiological, clinical, laboratory and radiological characteristics and treatment, and outcomes data were extracted from patients’ electronic medical records. The data were reviewed by a trained team of doctors in the hospital who recorded patients’ demographic data, laboratory findings, exposure history, comorbidities, symptoms and treatment measures.

Throat swab specimens were collected from all patients at admission, and 2019-nCoV ribonucleic acid was detected by real-time polymerase chain reaction within 3 hours, as in a previous study. Virus detection was repeated twice every 24 hours for 3 days.

Cardiac injury was diagnosed if the serum levels of troponin I (TNI) were above the 99th percentile of the upper reference limit (> 0.03 μg/L) using the Access AccuTnI+3 test (Beckman Coulter Inc., Brea, CA, USA). The highest level of TNI was used to evaluate the degree of myocardial injury. Unless otherwise specified, all values are the first data after admission, and if the index was measured more than twice, we chose the highest value for analysis. Symptoms, ARDS and intensive care unit (ICU) admission were recorded, and we defined ARDS severity according to the international guidelines for community-acquired pneumonia^11^. Preexisting cardiac conditions were defined as congestive heart failure, diabetes, arrhythmia or ischemic heart disease and hypertension.

### Statistical Analysis

Continuous variables were expressed as mean ± standard deviation for normally distributed data or as median (interquartile range, IQR) for skewed distributions. Frequency data were presented as proportions. We compared continuous variables using Student’s t test or the Mann–Whitney U test when appropriate, whereas differences in categorical variables were assessed using the Chi-square test or Fisher’s exact test.

All analyses were performed using SPSS 25.0 (IBM Corp. Armonk, NY, USA). Statistical charts were created using GraphPad Prism 7 software (GraphPad, San Diego, CA, USA), and a two-tailed *p*-value < 0.05 was considered statistically significant.

## Results

### 1. Epidemiological features of 2019-nCoV-infected patients with cardiac Injury

A total of 291 patients from Guangzhou Eighth People’s Hospital were included in the final analysis, and 15/219 patients (5.2%) had cardiac injury. Patients with cardiac injury had a higher mean age than these without cardiac injury. The median age was 65 years (interquartile range, 55–72), and 11/15 (73.3%) were men. Patients with cardiac injury had a higher proportion of preexisting cardiac conditions such as hypertension (46.6%) and coronary heart disease (20%). The epidemiological characteristics of the study participants are presented in Table 1.

**Table 1.** Epidemiological features of 2019-nCoV-infected patients with cardiac injury.

| Variables | All patients<br>(n=291) | With<br>cardiac<br>injury(n=15) | Without<br>cardiac injury(n=276) | *P* |
| --- | --- | --- | --- | --- |
| Age (Y), | 48.1 (34-62 ) | 65(55-72) | 47(33-61) | 0.001* |
| Male, N (%) | 133(45.7) | 11 ( 73.3 ) | 122 ( 44.2 ) | 0.03* |
| Exposure history in Wuhan | 149(51.2) | 8(53.3) | 141(51.1) | 1 |
| Preexisting condition, N (%) |  |  |  |  |
| Diabetes | 22(7.6) | 3(20) | 19(6.9) | 0.09 |
| Hypertesion | 54(18.5) | 7(46.7) | 47(17) | 0.01* |
| CHD | 12(4.1) | 3(20) | 9(3.3) | 0.02* |
| Heart failure | 1(0.3) | 1(6.7) | 0(0) | 0.051 |
| Arrhythmia | 2(0.7) | 1(6.7) | 1(0.4) | 1.006 |

CHD :coronary heart disease.Values are numbers (percentages) unless stated otherwise
\**P*<0.05

### 2. Clinical features and laboratory findings of 2019-nCoV-infected patients with cardiac injury

2019-nCoV-infected patients with or without cardiac injury had similar symptoms. The most common symptoms at illness onset were fever (n = 11, 73.3%), cough (n = 7, 46.7%), headache or fatigue (n = 5, 33.3%) and dyspnea (n = 4, 26.6%). No patients with cardiac injury complained of chest pain and palpitation, and the values for most of the laboratory results were within the normal ranges. However, patients with cardiac injury had higher systolic blood pressure (132 mmHg vs 124 mmHg), White blood cell count(5.6 × 10^9^/L vs 5.2× 10^9^/L, neutrophil count(4.1×10^9^/L vs 3.0.×10^9^/L, troponin I(0.07 ug/L vs 0.003 ug/L), brain natriuretic peptide(245.5 pg/mL vs 18.5 pg/mL), D-dimer(2430 mg/L vs 1090 mg/L) and lower lymphocyte counts (1.1 × 10^9^/L vs 1.6 × 10^9^/L) and platelet counts (165 × 10^9^/L vs 208 × 10^9^/L) vs patients without cardiac injury, respectively. However, in all patients with cardiac injury, C-reactive protein levels were elevated, and patients had bilateral infiltrates on chest X-ray. The clinical features and selected laboratory findings of the study participants are presented in Table 2.

**Table 2.** Clinical features and laboratory findings of 2019-nCoV-infected patients with cardiac injury.

| Variables | Normal | All patients | With cardiac | Without cardiac | P |
| --- | --- | --- | --- | --- | --- |
|  | range | (n=291) | injury(n=15) | injury(n=276) |  |
| Cough, N (%) | - | 164(56.4) | 7(46.7) | 157(56.9) | 0.44 |
| Fever, N (%) | - | 193(66.3) | 11(73.3) | 182(65.9) | 0.78 |
| Headache/Fatigue, N (%) | - | 53(18.2) | 5(33.3) | 48(17.4) | 0.16 |
| Dyspnea, N (%) | - | 33(11.3) | 4( 26.7 ) | 29( 11 ) | 0.08 |
| Chest pain, N (%) | - | 1(0.3) | 0(0) | 1(0.4) | 1 |
| palpitation, N (%) | - | 3(1) | 0(0) | 3(1.1) | 1 |
| Heart rate ( bpm ) | 60-100 | 84(78-92) | 81(72-96) | 84(78-92) | 0.44 |
| Highest<br>temperature (°C) | 36.3-37.3 | 36.9(36.6-37.5) | 37.5(36.5-38.8) | 36.9(36.6-37.4) | 0.06 |
| Systolic<br>pressure(mmHg) | 90-139 | 124(117-136) | 132(125-143) | 124(116-135) | 0.017* |
| White blood cell count<br>(10E9/L) | 4-10 | 5.22(4.15-6.45) | 5.62(5.14-11.51) | 5.15(4.1-6.39) | 0.023* |
| Neutrophil<br>count<br>(10E9/L) | 1.8-6.3 | 3.03(2.2-4.0) | 4.06(3.7-10.1) | 2.96(2.1-3.9) | <0.001* |
| Lymphocytes<br>count<br>(10E9/L) | 1.1-3.2 | 1.4(1.1-2.0) | 0.98(0.8-1.2) | 1.5(1.1-2.0) | 0.001* |
| Hemoglobin, g/L | 113-151 | 135(123-147) | 122(111-141) | 135(124-147) | 0.053 |
| Platelets count (10E9/L) | 100-300 | 206(159-248) | 165(137-188) | 209(160-250) | 0.022* |
| C reactive protein (>10<br>mg/L,N%)* | <10 | 112(38.5) | 15(100) | 97(35) | 0.0001* |
| Troponin I, (ug/L) | <0.03 | 0.004(0.001-0.008) | 0.07(0.04-0.23) | 0.003(0.001-0.006) | <0.001* |
| Brain natriuretic peptide<br>(pg/mL) | 0-100 | 35.5(12.8-111.8) | 245.5(42.5-475.5) | 18.5(9.3-49.8) | <0.001* |
| Creatinine, µmol/L | 59~104 | 61.3(49.8-76.5) | 68.5(57.8-86.3) | 60.8(49-75.6) | 0.05 |
| Aspartate<br>aminotransferase (U/L) | 13-35 | 18.6(14.9-26.6) | 23.8(18.6-40.9) | 18.2(14.5-25.1) | 0.25 |
| Alanine<br>aminotransferase (U/L) | 7-40 | 22.1(14.3-34.5) | 23.4(13.2-47.8) | 22.05(14.3-34.4) | 0.40 |
| Creatine kinase (U/L) | 50-310 | 53.5(38-80.25) | 51(38-114) | 54(38-80) | 0.63 |
| D-dimer(mg/L) | <1000 | 1100(720-1700) | 2430(1090-3750) | 1090(700-1640) | 0.002* |
| Blood Oxygen Saturation | >94% | 98(97-98.8) | 97.9(97-99.1) | 98(97-98.8) | 0.07 |
| Bilateral involvement on<br>chest radiographs, N (%) | - | 242(83.1) | 15(100) | 227(82.2) | 0.08 |

Values are numbers / medians (percentages or interquartile ranges) unless stated
otherwise.\* $P<0,05$

### 3. Treatments and outcomes of 2019-nCoV-infected patients with cardiac Injury

Complications included ARDS (20%) and severe pneumonia (73.3%), and these were common in patients with cardiac injury. A greater proportion of patients with cardiac injury required tracheal cannula(46.7% vs 0.4%, P<0.0001), invasive mechanical ventilation (53.3% vs 8.3%; p < 0.0001), continuous renal replacement therapy (33.3% vs 0%; p < 0.0001), extracorporeal membrane oxygenation (26.7% vs 0%; p < 0.0001), vasopressor therapy (20% vs 0.4%; p < 0.0001) and admission to the ICU (73.3% vs 5.4%; p < 0.0001) compared with patients without cardiac injury, respectively. One patient with cardiac injury died during the study. Treatments and outcomes of 2019-nCoV-infected patients with cardiac injury are shown in Table 3.

**Table 3.** Treatments and outcomes of 2019-nCoV-infected patients with cardiac injury.

| Treatments and outcomes | All patients<br>(n=291) | With cardiac<br>injury(n=15) | Without cardiac<br>injury(n=276) | p |
| --- | --- | --- | --- | --- |
| Tracheal cannula,N(%) | 8(2.7) | 7(46.7) | 1(0.4) | <0.00* |
| IMV,N(%) | 31(10.7) | 8(53.3) | 23(8.3) | <0.00* |
| Vasopressor therapy,N(%) | 4(1.4) | 3(20) | 1(0.4) | <0.00* |
| CRRT,N(%) | 5(1.7) | 5(33.3) | 0 ( 0 ) | <0.00* |
| ECMO,N(%) | 4(1.4) | 4(26.7) | 0(0) | <0.00* |
| ARDS,N(%) | 3(1) | 3(20) | 0(0) | <0.00* |
| Severe pneumonia,N(%) | 29(9.9) | 11(73.3) | 18(6.5) | <0.00* |
| Admission to ICU,N(%) | 26 ( 8.9 ) | 11 ( 73.3 ) | 15 ( 5.4 ) | <0.00* |
| Death,N(%) | 1 ( 0.3 ) | 1 ( 6.7 ) | 0 ( 0 ) | 0.05 |

3 CRRT :continuous renal replacement therapy; ECMO: extracorporeal membrane oxygenation.ICU: intensive care 4 unit. ARDS : acute respiratory distress syndrome. IMV:Invasive mechanical ventilation. Values are numbers 5 (percentages) of patients.\* $P<0.0001$

## Disccussion

To our knowledge, this is one of largest studies to systematically investigate cardiac injury in hospitalized patients with 2019-nCoV infection. In this retrospective single-centre study, a small proportion (5.2%) of patients with 2019-nCoV infection were diagnosed with cardiac injury. Our results showed that 2019-nCoV-infected patients with cardiac injury had worse clinical outcomes compared with patients without cardiac injury.

Cardiac injury is common with infections caused by influenza virus, Coxsackie virus and other viruses, although viral-induced cardiac damage is relatively mild ^3^. Even the fatal coronaviruses that caused middle east respiratory syndrome and sudden acute respiratory syndrome resulted in minimal heart damage e^4, 5^. 2019-nCoV is a new coronavirus, and it is unclear whether it causes serious heart damage. Recent studies have yielded inconsistent results. Xu et al found that there was no obvious heart damage in autopsy examinations of patients with 2019-nCoV-induced pneumonia^6^. However, other studies found that patients with 2019-nCoV-induced pneumonia also developed cardiac injury ^7 8 9^. To ensure the accuracy of our cardiac injury evaluation, we chose TNI as the only evaluation index. As a gold standard to evaluate cardiac injury, TNI is more accurate than creatine kinase-MB and electrocardiography. Our study found that a small proportion of 2019-nCoV-infected patients (5.2%) developed cardiac injury, which is a lower rate than in previous studies ^9^. This result may be related to using TNI as the only evaluation index in this study.2019-nCoV-infected patients with cardiac injury in our study were usually older (median age: 62 y) and often had pre-existing heart disease (hypertension, coronary heart disease). Furthermore, older patients with chronic disease are more likely to be critically ill ^9^. Our results showed that patients with a history of heart disease are at increased risk of serious illness or death if they are infected with 2019-nCoV.

The mechanism of cardiac injury in patients infected with 2019-nCoV is unclear. In a recent study, autopsy examinations failed to detect 2019-nCoV in heart tissue, and the authors found no other substantial damage to the heart tissue ^6^. However, increased numbers of studies have indicated an association between 2019-nCoV infection and cardiac injury^, 12^. To explain these inconsistencies, severe infection, hypoxia and mechanical ventilation settings may be associated with cardiac injury, and these common conditions in patients infected with 2019-nCoV may partly explain the heart damage. It is also very possible that the viral invasion of 2019-nCoV to the heart does not occur directly, but indirectly, via the inflammatory response. Severe 2019-nCoV infection may trigger an exaggerated immune response. This finding was confirmed by autopsy results ^6^, which showed that overactivation of T cells accounts, in part, for the severe immune injury. Another study showed that 2019-nCoV-infected patients admitted to the ICU have higher cytokine levels (interleukin 2, interleukin 7 and other cytokines) compared with patients not admitted to the ICU ^13^. According to these findings, we hypothesize that 2019-nCoV may promote cardiac injury by an inflammatory reaction, but this hypothesis must be explored further.

Our study showed that during COVID-19 epidemics, cardiac injury was associated with clinical outcomes for 2019-nCoV-infected patients. The proportion of patients requiring non-invasive ventilator support and extracorporeal membrane oxygen, admission to ICU and developing ARDS was significantly higher in patients with cardiac injury compared with patients without cardiac injury. Cardiac injury is a potential indicator of risk stratification for 2019-nCoV-infected patients.

### Limitations

There are several limitations in our study. First, this study was a single-centre study involving a small number of patients, and evaluating more medical records is needed to support our conclusions. Second, because our patients were from Guangzhou City, only, different clinical features of patients with heart injury from other geographic areas may be found in future studies. Third, of the 291 patients, some were still hospitalized at submission of this manuscript. Therefore, it is difficult to assess patients’ long-term prognosis, and it is necessary to continue to observe the natural history of the disease.

## Conclusions

Cardiac injury is a common condition among patients infected with 2019-nCoV. Compared with patients without cardiac injury, the clinical outcomes of patients with cardiac injury are relatively worse. Cardiac injury is a potential prognostic risk indicator for patients infected with 2019-nCoV.

## Data Availability

All data are available when requested.

## Acknowledgements

The authors had full access to all of the data in the study and take responsibility for the integrity of the data and the accuracy of the data analysis. Study concept and design: Youbin Liu, Zhongwei Hu, Zhihui Qin and Jinglong Li. Acquisition of data: Dehui Liu, Huafeng Song, Chunlin Chen, Mingfang Lv, Xing Pei, Jinglong Li. Analysis and interpretation of data: Dehui Liu, Huafeng Song, Chunlin Chen, Mingfang Lv, Xing Pei, Jinglong Li. Drafting of the manuscript: Youbin Liu and Jinglong Li. Critical revision of the manuscript for important intellectual content: Youbin Liu, Dehui Liu, and Jinglong Li. Statistical analysis: Youbin Liu and Jinglong Li. Administrative, technical, or material support: Dehui Liu, Huafeng Song, Chunlin Chen, Mingfang Lv, Xing Pei, Jinglong Li. Supervision: Youbin Liu, Zhongwei Hu, Zhihui Qin and Jinglong Li.

